# COVID-19 lockdown: if, when and how

**DOI:** 10.1101/2020.06.20.20136325

**Authors:** Lía Mayorga, Clara García Samartino, Gabriel Flores, Sofía Masuelli, María V. Sánchez, Cristián G. Sánchez, Luis S. Mayorga

## Abstract

**Background:** With the lack of an effective SARS-CoV-2 vaccine, mathematical modeling has stepped up in the COVID-19 management to guide non-pharmaceutical intervention (NPI) policies. Complete lockdown has been characterized as the most powerful strategy for the epidemic; anyhow, it is associated with undeniable negative consequences. Not aware that global panic could make countries adopt premature and lengthy lockdowns, previous studies only warned about the inefficacy of late quarantine sets. Therefore, we proposed ourselves to find the optimal timing and lasting for COVID-19 suppressive measures.

**Methods:** We used our previously elaborated compartmental SEIR (Susceptible-Exposed-Infected-Recovered) model to scan different timings for lockdown set and various lockdown lengths under different reproduction number (R_0_) scenarios. We explored healthcare parameters focusing on ICU occupation and deaths since they condition the sanitary system and reflect the severity of the epidemic.

**Results:** The timing for the lockdown trigger varies according to the original R_0_ and has great impact on ICU usage and fatalities. The less the R_0_ the later the lockdown should be for it to be effective. The lockdown length is also something to consider. Too short lockdowns (∼15 days) have minimal effect on healthcare parameters, but too long quarantines (>45 days) do not benefit healthcare parameters proportionally when compared to more reasonable 30 to 45-day lockdowns. We explored the outcome of the combination of a 45-day lockdown followed by strict mitigation measures sustained in time, and interestingly, it outperformed the lengthy quarantine. Additionally, we show that if strict mitigation actions were to be installed from the very beginning of the epidemic, lockdown would not benefit substantially regarding healthcare parameters.

**Conclusion:** Lockdown set *timing and lasting* are non-trivial variables to COVID-19 management.

## BACKGROUND

COVID-19 pandemic has changed our lives since its outbreak by the end of 2019. The world is still struggling to control this catastrophe. Scientists from different areas of expertise are working non-stop looking for a treatment, a vaccine, or the optimal public health management. Without the possibility of immunization in the immediate future[1], non-pharmaceutical interventions (NPI) remain the most effective action. In the guidance of these NPIs mathematical modeling of the epidemic has been a critical factor.

NPIs can be divided into mitigation and suppression strategies. Mitigation criteria involve hygienic recommendations, case-isolation, general social distancing, banning of public gatherings, college closures, etc. Suppression measures include complete lockdown or quarantine of the whole population except for essential activities, intending to lower the reproduction number to ≤1 [2].

It is clear that the most potent strategy today against SARS-CoV-2 is complete lockdown, still, this is not without collateral damage. Most evidently, the economy has been hurt worldwide impacting as well directly in human health (increase in poverty, stress, lack of preventive medical interventions)[3,4]. The other undeniable consequences of lockdown are of psychological nature due to confinement, as addictions, domestic violence, increase in obesity, deterioration of pre-existing psychiatric illnesses, disturbance in the education especially in children[5–8], etc.

Mathematical modeling of the epidemic has been at the service of public health strategies early in the pandemic and has supported the use of lockdown as a necessary measure to overcome COVID-19 [2,9]. However, not aware that global fear could make countries adopt an extremely premature suppressive policy, none have alerted of the importance of the correct lockdown timing set.

In response to the different strategies adopted world round, we asked ourselves if the timing and lasting of the different NPIs were critical variables to take into account when embracing them. Furthermore, we proposed ourselves to analyze the optimal setting of different stages of NPIs and the possibility of combining them to achieve the best possible outcomes.

## METHODS

We based this paper on our previously elaborated SEIR (Susceptible-Exposed-Infected-Recovered) model for COVID-19 epidemics that incorporates specific compartments. We modeled asymptomatic or very mildly affected individuals as a subset of the infectious compartment and integrated healthcare burden parameters[10]. In the supplementary material (supplementary figures and tables), we expose the compartments and parameters that we used in our model. Furthermore, we share the set of differential equations that give rise to the model.

We considered an infective injection of 100 symptomatic cases/1 million inhabitants on day 0 for the modeled scans.

To facilitate the use of the model, the simulation was programmed in the free software COPASI (copasi.org), and the corresponding file is provided as supplementary material.

## RESULTS

### When to start lockdown? Under which primary conditions

If a single lockdown was raised as the strategy, when would it be more beneficial to start it? Is a premature or late quarantine effective as far as healthcare parameters? Under which basic reproduction conditions (R_0)_ would it be more convenient to trigger lockdown?

To answer these questions, we scanned the set of a single 45-day lockdown at different times during the epidemic progression in an R_0_=2.3 scenario. In other words, at different times, the R_0_ was switched from 2.3 to 1 for 45 days and then raised to 2.3 until the end of the simulation. The results are shown in video 1 (Supplementary material, video 1) and Figure 1. If the lockdown is initiated too early, its only effect is to delay the epidemic’s onset without affecting the final fatality number nor the maximal ICU occupancy (Fig. 1A). According to our scan, to minimize the ICU requirements over time, quarantine should start at day 87 (Fig. 1B). To reduce deaths, lockdown should be triggered on day 105 (Fig. 1C). Finally, as expected, a late start has no effect on the epidemic consequences (Figure 1D). It is worth noticing that the least fatalities do not match with the least ICU occupation, this is because the number of deaths accumulate in time and ICU holding changes over time according to the incorporation and release of patients to the healthcare system. One would obviously prefer to minimize deaths. Even so, the capacity of the local healthcare system regarding ICU should be taken into account when deciding the best option for the local scenario.

**Figure 1:**
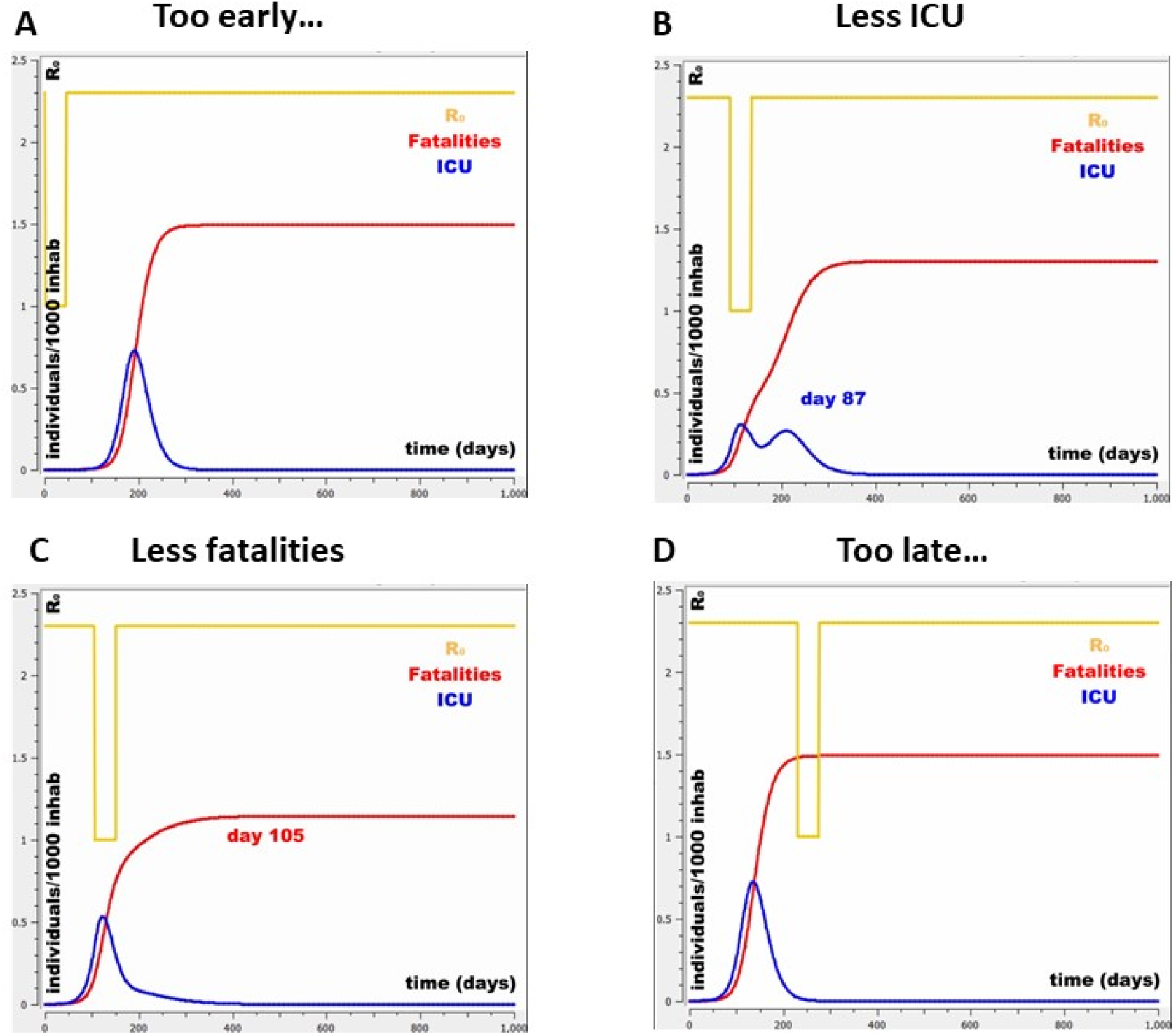
Quarantine set at different times in a R_0=_2.3 scenario. **A** and **D** show too early and too late lockdowns without any impact on healthcare parameters (ICU and deaths). **B** and **C** show effective lockdown settings regarding ICU occupancy (B) and fatalities (C).

It is evident that the timing for quarantine set is vital for a better outcome. To explore the best lockdown starting times to minimize fatalities and ICU occupancy, the same scan was performed under different R_0_s. Considering R_0_=4 for a totally uncontrolled scenario (as observed in Europe in March 2020[11]), R_0_=2.3 as considered in the first instance of the epidemic in Wuhan, China[12], R_0_=1.5 as strict mitigation measures and R_0_=1 as complete lockdown.

Figure 2 shows the maximum ICU usage and fatalities, according to when the lockdown is started. The lower the basal R_0_ is, the later the effective lockdown should be, and the fewer deaths and ICU use is observed. Furthermore, the lower the R_0,_ the less the slope of the curve and the more flexible the period for which an effective lockdown could be triggered. Additionally, this analysis shows that if strict mitigation measures could be sustained in time from the beginning of the epidemic (initial R_0_=1.5) lockdown would not change deaths nor ICU requirements substantially.

**Figure 2:**
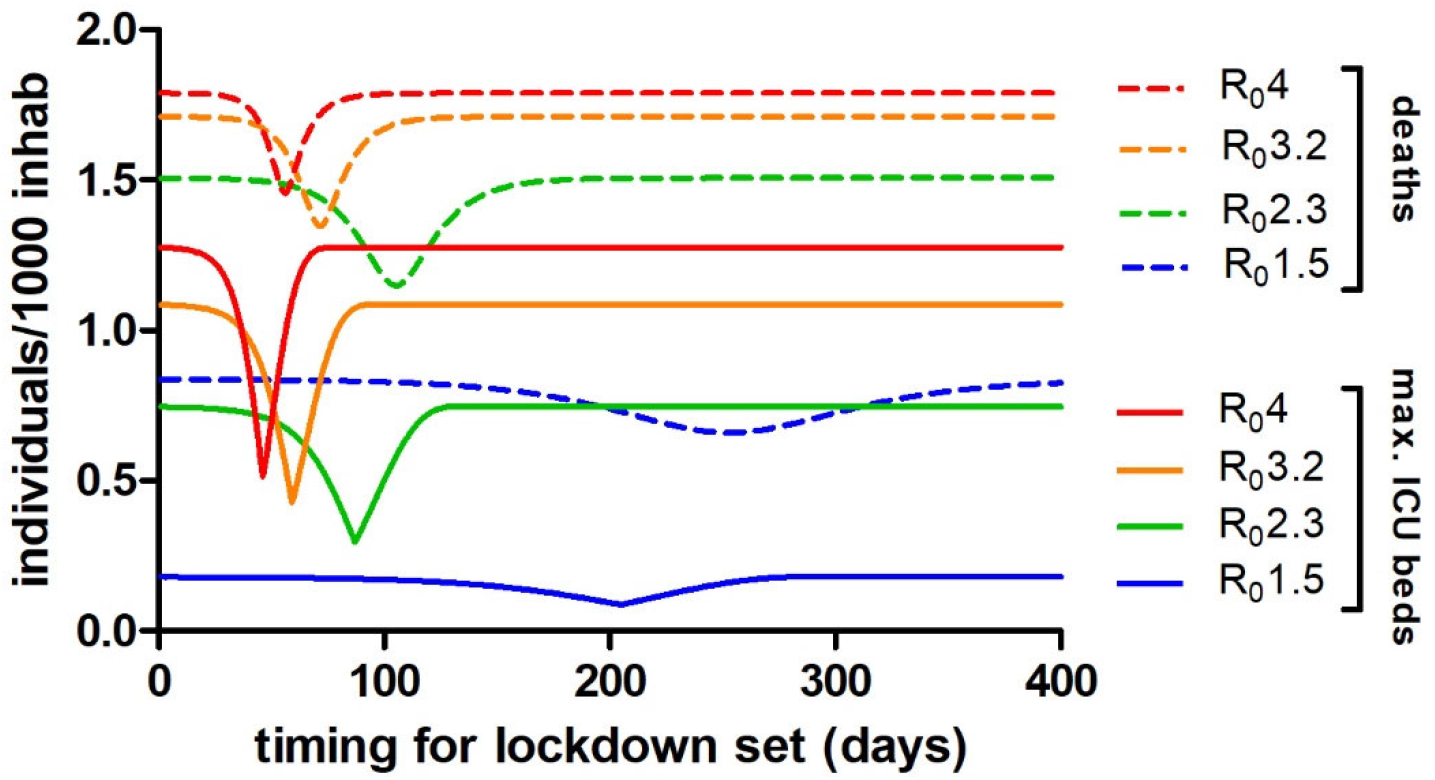
Fatalities and maximum ICU occupancy according to the R_0_ and lockdown set timing (a single 45-day lockdown was used for this scan).

### How long should lockdown last

Next, we wanted to explore how long the quarantine should last for it to be efficient. With that aim, we scanned different lockdown periods and evaluated the impact on deaths and ICU tenancy for an R_0_=2.3 scenario. As expected, the longer the lockdown, the fewer deaths and ICU requirements. Nonetheless, the advantage in healthcare parameters and quarantine lasting is not linear. The benefit in healthcare variables increases significantly when comparing a 15-day with a 30-day or 45-day lockdown. Quarantines longer than that do not benefit much more regarding deaths nor ICU occupation (Figure 3)

**Figure 3:**
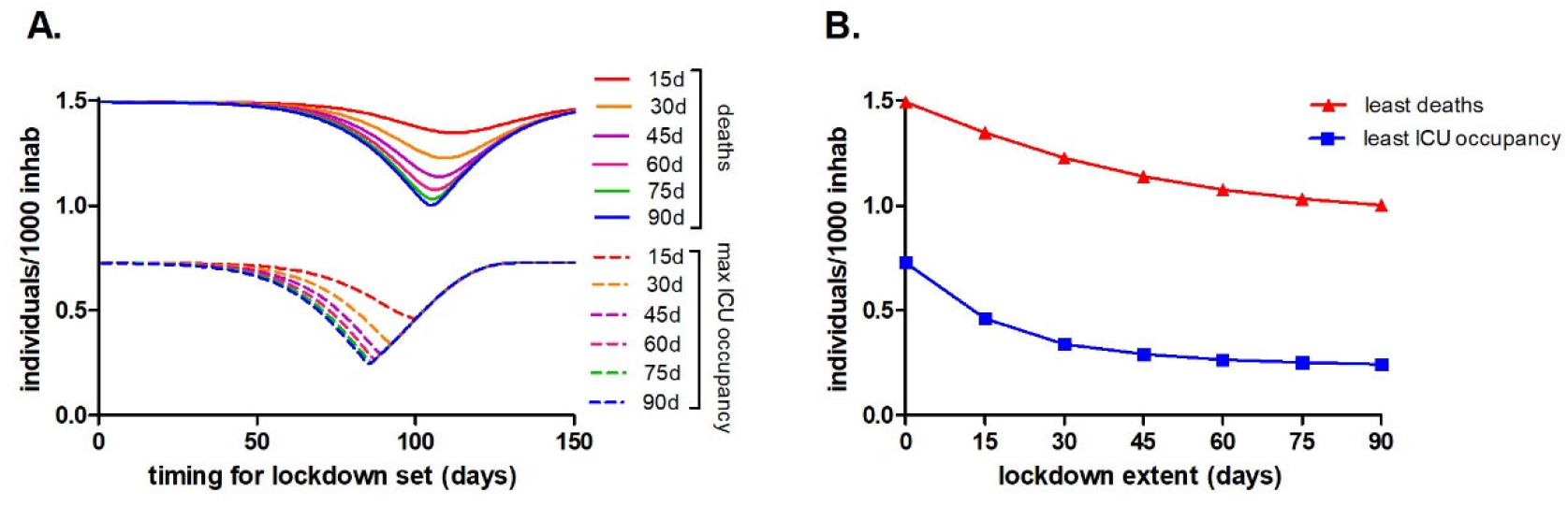
**A**. Different lockdown periods set at various timings and their impact on healthcare burden parameters (deaths and ICU occupation) in a R_0_=2.3 scenario. **B**. Different lockdown periods and their best efficacy in lowering deaths and ICU occupation in the same R_0_=2.3 scenario.

### Combining strategies (lockdown + strict mitigation)

A 3-month lockdown, apart from untenable, would be extremely demanding due to the social and economic impact. Furthermore, the benefit is not significant compared to a 45-day confinement period. Hence, we ventured to see if we could equal the healthcare parameters obtained with the lengthy lockdown combining a 45-day quarantine followed by strict mitigation strategies (R_0_=1.5) for a prolonged period (270 days) awaiting the arrival of an effective vaccine or heard immunization.

In video 2 (Supplementary material, video 2) and Figure 4 we explore when this combined strategy would be optimal to be set according to the aimed healthcare parameters. Day 73 and day 100 would be the optimal quarantine time sets to minimize ICU and deaths, respectively. Again, lockdown timing shows to be crucial to obtain the best outcome.

**Figure 4:**
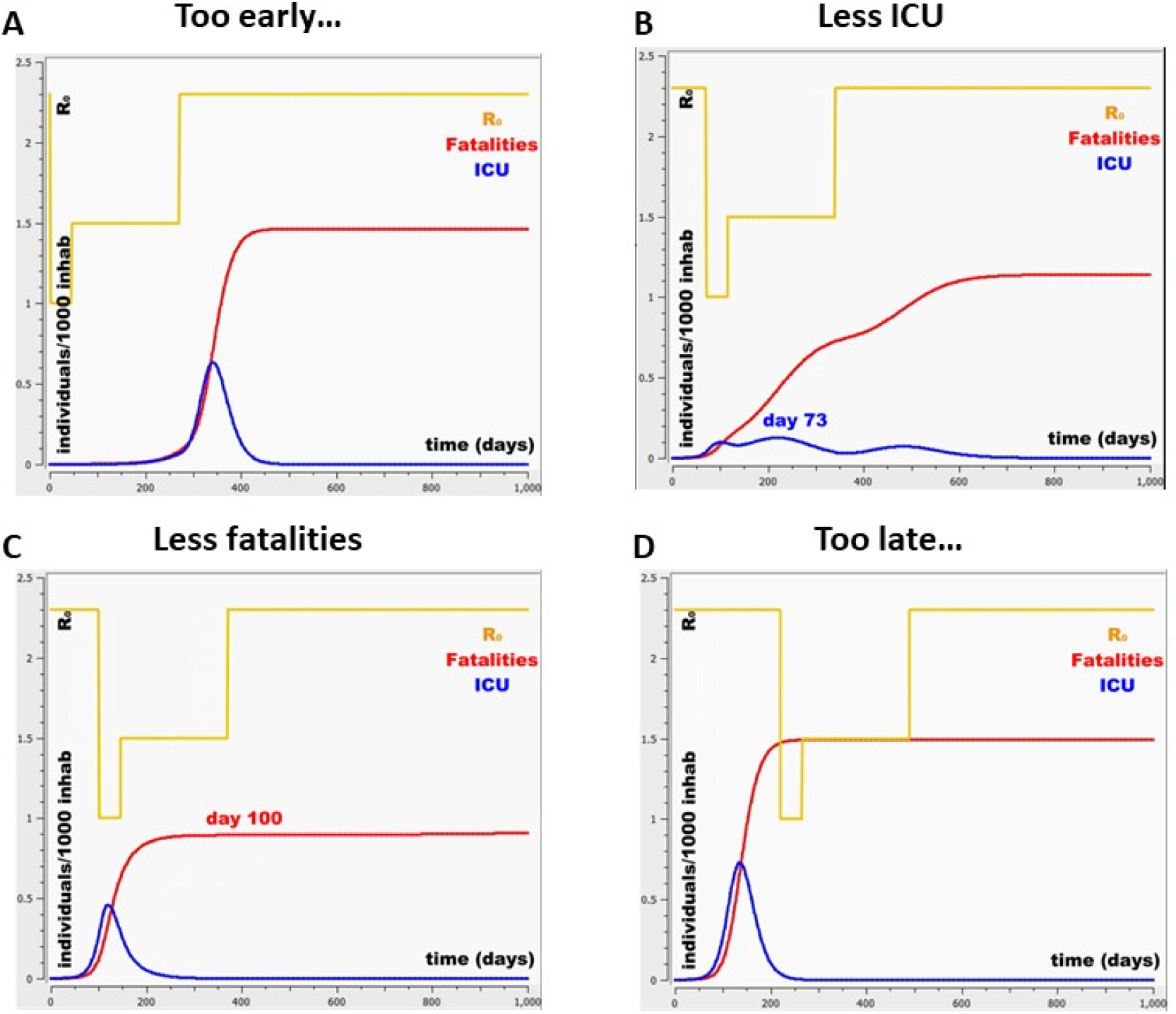
Combined lockdown strategy in a R_0_=2.3 scenario (45-day lockdown plus strict mitigation R_0_=1.5 over 270 days) set at different times. **A** and **D** show too early and too late lockdowns with no impact on healthcare parameters (ICU and deaths). **B** and **C** show effective lockdown settings regarding ICU occupancy (B) and fatalities (C)

When comparing this combined strategy with the 90-day complete lockdown, the former outperformed the latter not only in the number of deaths and ICU usage but also, the time for which it is optimal to set quarantine is more flexible (Figure 5)

**Figure 5:**
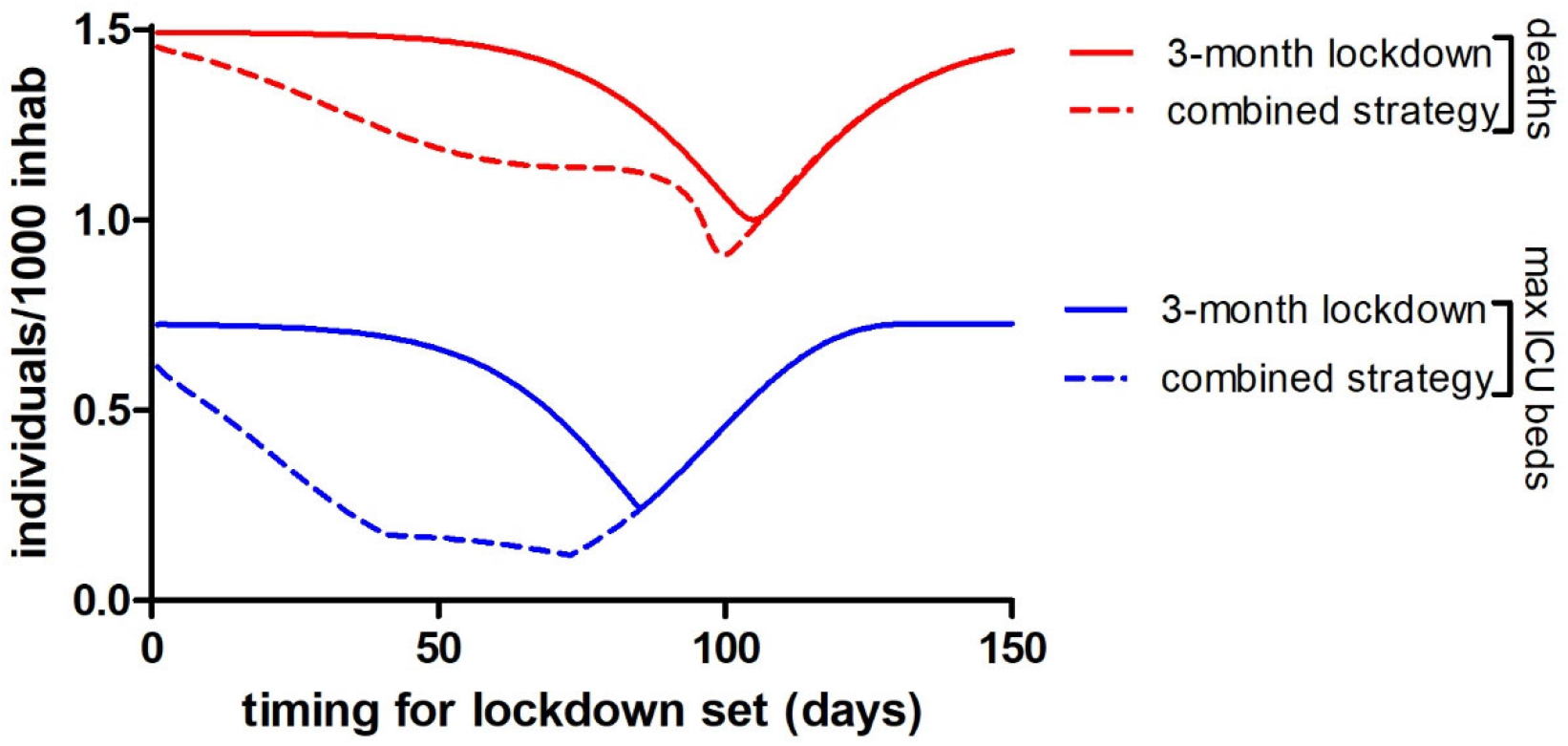
ICU occupation and COVID-19 deaths according to lockdown timing set in a R_0_=2.3 situation using a 3-month lockdown or a combined strategy= 45-day lockdown followed by strict mitigation measures (R_0_=1.5) for 270 days. The combined strategy benefits fatalities and healthcare burden and also shows a more flexible period to set a successful quarantine.

### Defining an objective lockdown trigger

It is easy to select the best day to start a lockdown after performing a scan in a simulation. However, the same decision is exceedingly difficult in the real-life epidemic situation. When to start a lockdown depends on a large and complex set of factors, including R_0_ and the present state of the epidemic. It is impossible to determine when day 1 is because the outbreak can be missed at the beginning due to the mild symptoms of the majority and the injection of infectious cases may vary according to different circumstances.

As an exercise, and being aware of the oversimplifications implied, we searched for objective parameters other than “days” that could help determine *when* to start the strict confinement periods. We scanned a range of R_0_ associated to their optimal lockdown trigger day (regarding ICU occupation and deaths) and explored the number of hospitalized patients in ICU at that point. Interestingly, the ICU occupation level at which a successful quarantine should be triggered is similar in all R_0_>2. Considering that in R_0_ <2, lockdown is questionable, we thought that ICU occupation level was the best impartial parameter to ponder as a lockdown trigger. Notably, the level of ICU patients at lockdown set to favor the least ICU usage is practically the same for all R_0_s. Using a 45-day quarantine strategy, for R_0_ higher than 2, to favor the least ICU occupation, the trigger for lockdown should be around 100 occupied ICU beds/1million inhabitants. To favor the least mortality, the trigger should be an occupation of about 400 ICU beds/1 million inhabitants. (Figure 6)

**Figure 6:**
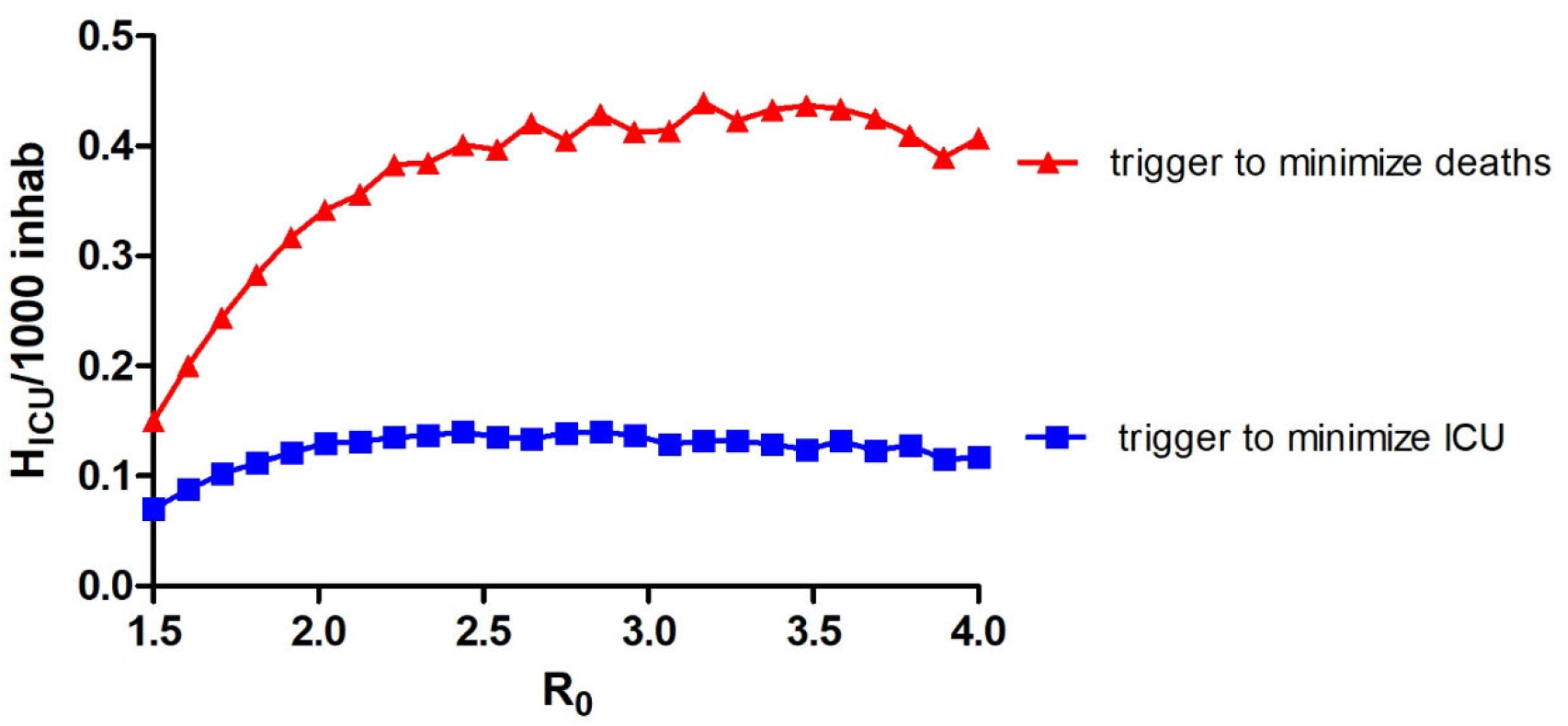
Defining an objective lockdown trigger. To determine the timing to set suppressive actions, we looked for objective healthcare burden parameters. Hospitalized patients requiring ICU(H_ICU_) can be used to mark the lockdown trigger. Using a 45-day lockdown strategy, for R_0_ >2, to favor the least ICU occupation, the trigger for lockdown should be ∼ 100 H_ICU_/1million inhabitants. To favor the least mortality, the trigger should be ∼ 400 H_ICU_/1 million inhabitants.

## CONCLUSIONS AND DISCUSSION

We have seen the world adopt a wide range of strategies in response to COVID-19 and all have had or are having different outcomes according to the local scenarios. Premature lockdowns have seemed to work in isolated places such as New Zealand where border’s irruption is difficult[13]. In most countries where borders are weak or large countries that need inner intercommunication to function, it seems that the epidemic will arrive sooner or later, and a premature lockdown will only postpone the arrival. In the optimistic view of an imminent vaccine this could be an ascertained strategy, but we will only know that in the months to arrive.

With the more skeptical assumption that a vaccine would only have a chance to be available after the end of 2020[1], the most logical strategy is to minimize the harm the epidemic can cause as it develops. For this, different non-pharmaceutical interventions (NPIs) are being adopted world round[2,11,14].

In this paper, we used a parametrized SEIR model to represent the COVID-19 epidemic and show that the timing for NPIs is not trivial to the healthcare outcomes. Too premature or too late lockdowns have *no* impact on deaths nor ICU occupation. We also evidenced that a higher ICU capacity would minimize deaths and shorten the epidemic period, so expanding the healthcare system could be a profitable but complicated strategy.

In addition, it is clear that mitigation measures cannot be released until an efficient vaccine arrives or until heard immunization is reached. Sustained and firm mitigation measures from the beginning could even avoid the need for a strict lockdown period and could be an alternative for areas with low ICU capacity.

The particular days to start a lockdown strategy to minimize harm are greatly influenced by the selected model and its parameters. Nevertheless, in most of the conceivable scenarios, the level of hospitalized patients in ICU at a specific time can be used as a reliable lockdown trigger.

The conclusion of this work supports that lockdown set *timing and lasting* are transcendent variables to COVID-19 management.

## Data Availability

The authors confirm that the data supporting the findings of this study are available within the article and its supplementary materials.
Data sharing: Code used in this study is available in the GitHub repository https://github.com/ihem-institute/SEIR_Mendoza.

## DECLARATIONS

### Data sharing

Code used in this study is available in the GitHub repository https://github.com/ihem-institute/SEIR_Mendoza

### Author approval

All authors have read and approved the manuscript.

### Author contributions

LSM and CGS conducted the research project. CGS was in charge of the mathematical model design and the implementation of it with the help of LSM and GF. LM chose the epidemiological compartments and variables with the help of CGSam, SM and MVS. LSM carried out lockdown scans using COPASI. LM, LSM and CGS wrote the manuscript and elaborated the figures. All authors have reviewed and approved the manuscript.

